# The “Hamburg Life After Cancer Program”: Development, Implementation and Evaluation of a Structured Survivorship Program - Study Protocol of a Hybrid Effectiveness-Implementation Study

**DOI:** 10.64898/2026.09.11.26362829

**Authors:** Hannah Führes, Julia von Grundherr, Franziska Wolters, Mareike Rutenkröger, Antonia Zapf, Ann-Kathrin Ozga, Annika Möhl, Otmar Kodalle, Holger Schulz, Carsten Bokemeyer, Anne Letsch, Isabelle Scholl, Marianne Sinn

**Author notes:** **Corresponding Author** Name: Hannah Führes University Medical Center Hamburg-Eppendorf Department of Medical Psychology Martinistr. 52, 20246 Hamburg, Germany. First author (Hannah Führes). Isabelle Scholl and Marianne Sinn share senior authorship.

## Abstract

**Background:** Advances in multimodal oncological treatment have improved survival, resulting in a growing population of cancer survivors. Yet, many survivors experience physical and psychosocial late and long-term effects after completing active cancer treatment, highlighting the need for follow-up care that extends beyond tumor surveillance. However, structured survivorship programs addressing survivors’ broader needs and evidence on their effectiveness and implementation remain limited.

**Methods:** This prospective, single-center, single-arm intervention study with an external control cohort aims to develop, implement, and systematically evaluate the *Hamburg Life After Cancer Program*. Applying a hybrid type 2 effectiveness-implementation design, the primary aim is to improve health-related quality of life (HRQoL) among program participants (n = 500), namely cancer survivors aged 18-65 years who completed primary treatment. Secondary objectives include improvements in various bio-psycho-social outcomes, as well as an exploratory comparison of HRQoL with an external control cohort receiving usual care (n = 250). Outcomes are assessed at baseline, 26 weeks and 52 weeks. Implementation is evaluated following a mixed-methods approach. The intervention comprises patient navigation, multidimensional assessment, survivorship consultations, an interdisciplinary survivorship board, individualized care plans, a survivorship handbook, educational sessions and peer-support. All study phases are guided by a participatory research approach involving patients and experts.

**Discussion:** Findings will contribute to the development of needs-based, feasible survivorship care models, improving HRQoL and providing holistic support for cancer survivors. The study design enables timely translation of the program into clinical practice and robust assessment of its effectiveness and implementation. However, the non-randomized design may limit causal inference and cannot fully exclude residual confounding or selection bias despite statistical adjustment. If shown to be effective and feasible, the program may serve as a standardized model for structured survivorship care in Germany.

**Trial Registration:** Clinical trial registration number: DRKS00035126, German Clinical Trial Register (date of registration: 27 August 2025).

## Background

As survival rates rise, cancer survivorship has internationally become an increasingly important topic in oncology and health care research. In the United States, more than 18 million individuals are currently living with a history of cancer, with projections indicating continued growth in the coming years [1]. Across Europe, over 23 million people are estimated to be cancer survivors, including approximately five million in Germany, reflecting similar demographic trends and highlighting the growing need for structured survivorship care [2, 3]. Given that the terms *cancer survivorship* and *cancer survivor* are used inconsistently in the literature and encompass varying conceptualizations [4-6], this study explicitly defines *cancer survivors* as individuals diagnosed with cancer who have completed primary treatment with curative intent, covering all phases of post primary treatment *cancer survivorship* from acute, extended and long-term survival [4, 5].

Although survival has improved due to advances in multimodal oncological treatment, these improvements are frequently accompanied by substantial physical, psychological, and social late and long-term effects, all of which can significantly reduce health-related quality of life (HRQoL) [7]. Common persistent symptoms include fatigue, pain, depression, fear of recurrence, cognitive impairments, sexual dysfunction, infertility, and neuropathy, while long-term risks such as cardiovascular disease and second primary cancers may emerge years or decades after treatment [7, 8]. Unhealthy lifestyle patterns, particularly overweight and obesity, remain highly prevalent and may further increase risks of recurrence or secondary malignancies [9, 10]. Further challenges, including social difficulties, financial strain, and occupational disruption, are commonly reported [7, 11], with social support recognized as a key protective factor throughout the survivorship continuum [12].

International health-policy agendas increasingly recognize cancer survivorship as a critical priority in modern cancer care. The recently published policy report by the European Cancer Organization for example calls for personalized long-term survivorship care plans with routine quality-of-life assessments and systematic collection of patient-reported outcome measures (PROMs) [13]. In Germany, the National Cancer Plan has acknowledged these complex needs and called for needs-based survivorship care models addressing physical, psychological, functional, and social dimensions [14]. Despite these recommendations, traditional follow-up care remains largely focused on tumor surveillance and recurrence detection. While essential, this approach often neglects survivors’ need for personalized, multi-professional care offering holistic support [15]. In Germany, survivorship care is fragmented, with limited structured assessments, insufficient coordination between providers, and few standardized approaches to long-term symptom management, prevention, empowerment, and health literacy [16]. Consequently, significant gaps remain in coordinated outpatient care after rehabilitation, highlighting the importance of collaboration with primary care physicians and other outpatient professionals as accessible and trusted points of contact [17, 18].

Modern survivorship care recognizes health literacy, empowerment, and self-management as essential components. Low health literacy, common among cancer patients and survivors, is associated with poorer quality of life and difficulties navigating the healthcare system [19]. Interventions such as patient navigation, tailored education, and cancer-specific digital tools have been shown to strengthen health literacy, enhance self-efficacy, reduce symptom burden - including fatigue, pain, and anxiety - and improve quality of life [20-22]. Digital tools such as cancer-specific apps have shown promise in supporting patients’ knowledge, motivation, and active participation in their care [22]. Patient navigation programs, widely implemented in the United States, have also been linked to higher patient satisfaction and fewer hospital admissions during follow-up [23].

Taken together, current evidence suggests that patient-centered survivorship care requires: (1) addressing individual needs and personal goals of survivors and their relatives regarding follow-up care and late or long-term effects; (2) strengthening health literacy, empowerment, self-management, and existing resources; (3) ensuring coordinated, continuous care through structured collaboration among primary care physicians, oncologists and further professions managing late and long-term effects (e.g., psycho-oncology, rehabilitation, nutrition, physiotherapy, social work); and (4) integrating community-based resources, including self-help organizations, counseling services, and health-promoting programs [16, 24].

Building on these recommendations, this study addresses the described care and research gaps by developing and implementing the *Hamburg Life After Cancer Program* as a structured, multi-component survivorship program for cancer survivors post primary cancer treatment in Germany. The study aims to evaluate the program’s effectiveness by assessing improvements in HRQoL (primary outcome) and secondary bio-psycho-social outcomes (e.g., anxiety, depression, empowerment, health literacy, self-efficacy, social support, nutrition), as well as its implementation with regard to acceptability, feasibility, patient-centeredness, barriers and facilitators.

## Methods

### Study design and setting

This study uses a type 2 hybrid effectiveness-implementation design, which allows for simultaneous evaluation of effectiveness and implementation [25]. The prospective, single-center, single-arm intervention study takes place at the University Cancer Center Hamburg (UCC Hamburg), one of 14 comprehensive cancer centers in Germany, embedded in the University Medical Center Hamburg-Eppendorf (UKE). An external control cohort receiving usual care is recruited at the Cancer Center Schleswig-Holstein in Kiel, part of the University Medical Center Schleswig-Holstein (UKSH). For further details on the study’s design and phases see figure 1.

**Figure 1.**
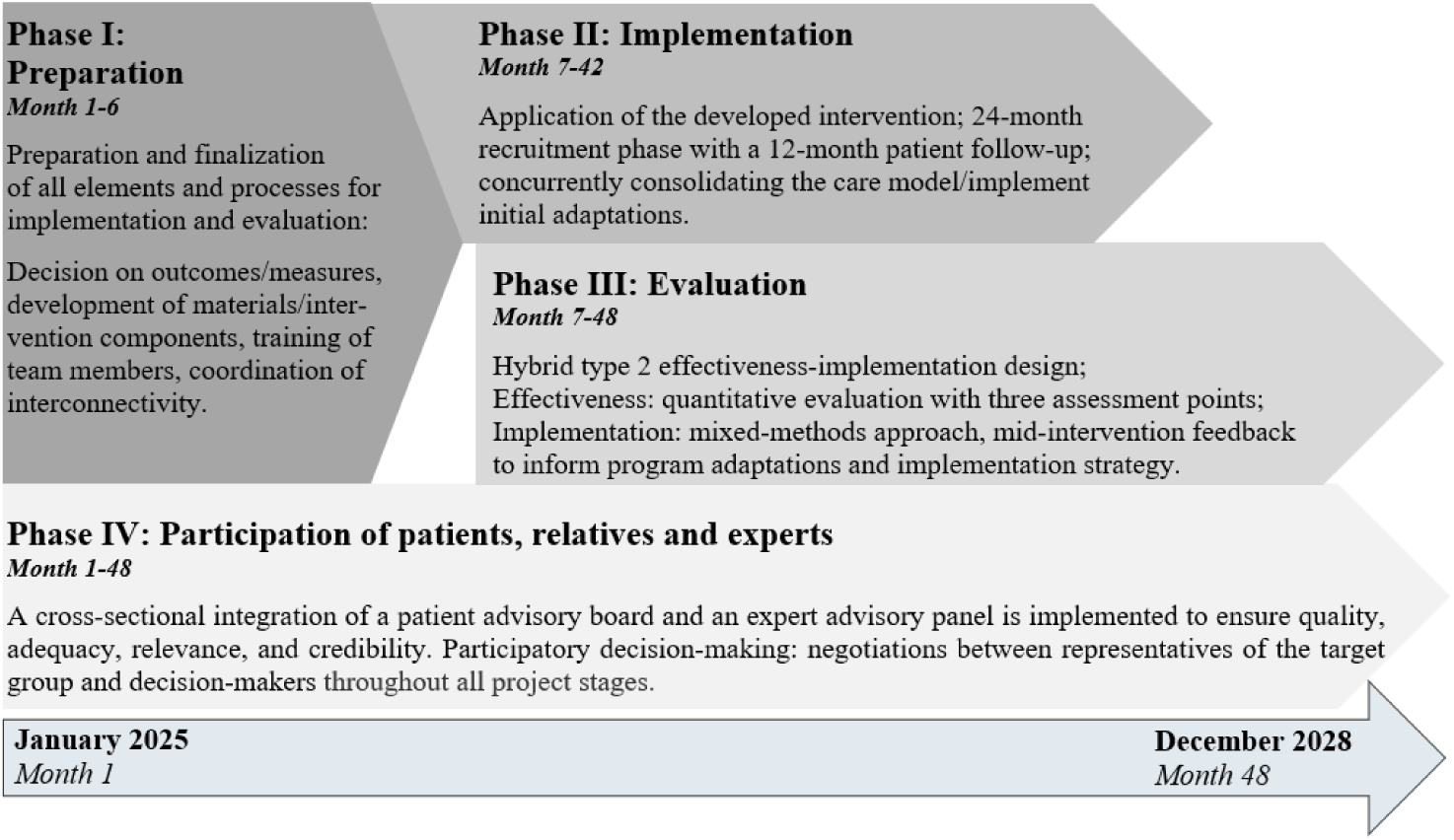
Overview of the study’s design and phases.

### Study population

Participants will be adult cancer survivors aged 18 to 65 years, diagnosed with any cancer entity (malignant neoplasms) classified according to the International Statistical Classification of Diseases and Related Health Problems (ICD-10, C00-C97) [26] at age 18 or older. In line with our definition of cancer survivors, eligible participants must have completed active cancer treatment - with the exception of adjuvant hormone therapy - with curative intent (i.e., post primary treatment). Accordingly, participation is eligible immediately after completion of active treatment, during tumor-specific follow-up (usually up to five years post-treatment) and long-term survivorship follow-up (more than five years post-treatment). Eligible participants must reside in the greater Hamburg metropolitan area for being able to attend at least one in-person appointment at the UCC Hamburg. Sufficient German language is required to understand and complete the study assessments. The control group is subject to the same inclusion criteria, except the area of residence.

### Intervention

The *Hamburg Life After Cancer Program* is a complex, multi-component survivorship care model developed in alignment with recommendations for needs-based survivorship care [16, 24, 27, 28]. Participants will complete the program over a period of one year. Figure 2 shows the participant pathway.

**Figure 2.**
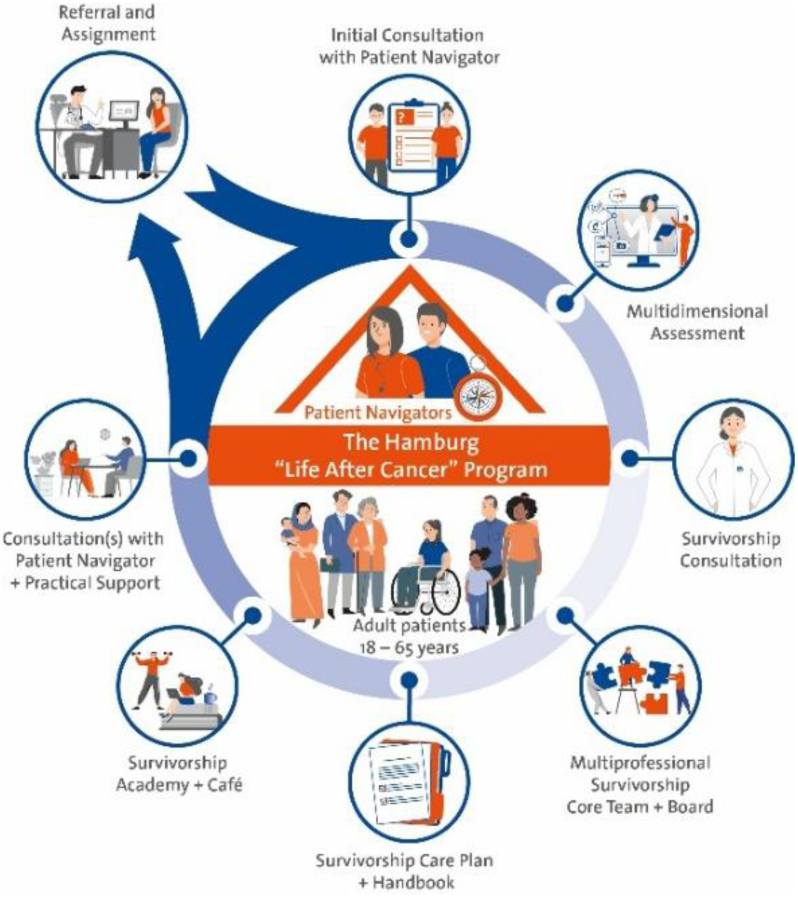
Overview of the structure and patient pathway in the Hamburg Life After Cancer Program.

The intervention comprises seven integrated components:

1. *Multidimensional Assessment:* Bio-psycho-social assessment using PROMs is conducted upon enrollment in the program and informs the subsequent components (survivorship navigation, consultation, and the multidisciplinary board). Based on international survivorship guidelines [24, 27, 28] the assessment screens for somatic late and long-term effects, psychological distress, social and occupational burden, lifestyle factors, health literacy, empowerment, personal resources and unmet needs (Tab. 1).
2. *Survivorship Navigator:* A trained patient navigator serves as the central point of contact, coordinates appointments, provides counseling, facilitates access to internal and external support services and delivers tailored information to accommodate individual health literacy needs. Participants may attend up to three patient navigator consultation appointments.
3. *Survivorship Consultation:* After the initial PROM assessment and consultation with the patient navigator, a consultation with an oncologist takes place. It includes clinical assessment, review of multidimensional assessment results, evaluation of late effects, guideline-based survivorship care recommendations and planning of further diagnostics. Further consultations are possible throughout the program, if needed.
4. *Survivorship Core Team and Multidisciplinary Board:* A weekly interdisciplinary board (including oncologists, patient navigators, psycho-oncologists, nutritionists, sport scientists, social workers, and primary care physicians) reviews each case and formulates individualized recommendations. The navigator represents patient perspectives to ensure patient-centered planning. Further specialized health care professionals (e.g., gynecologists) will be involved during the implementation period to complement the survivorship board.
5. *Survivorship Plan:* A structured survivorship plan is developed that summarizes clinical findings and provides individualized survivorship care recommendations based on the preceding program components (consultations and multidisciplinary board discussions). The plan is created using a shared decision-making approach and is accompanied by a plain-language summary to support patient empowerment and self-management.
6. *Survivorship Handbook:* A patient-centered survivorship handbook written in plain language provides survivorship care recommendations, lifestyle guidance, and information on regional and online resources. It is developed in close collaboration with a patient advisory board.
7. *Survivorship Academy and Café:* Monthly educational sessions are taking place to enhance health literacy and empowerment across modules such as nutrition, physical activity, psycho-oncology, work/family/social environment, and communication with healthcare professionals. A survivorship café offers peer and social support.

**Table 1.**
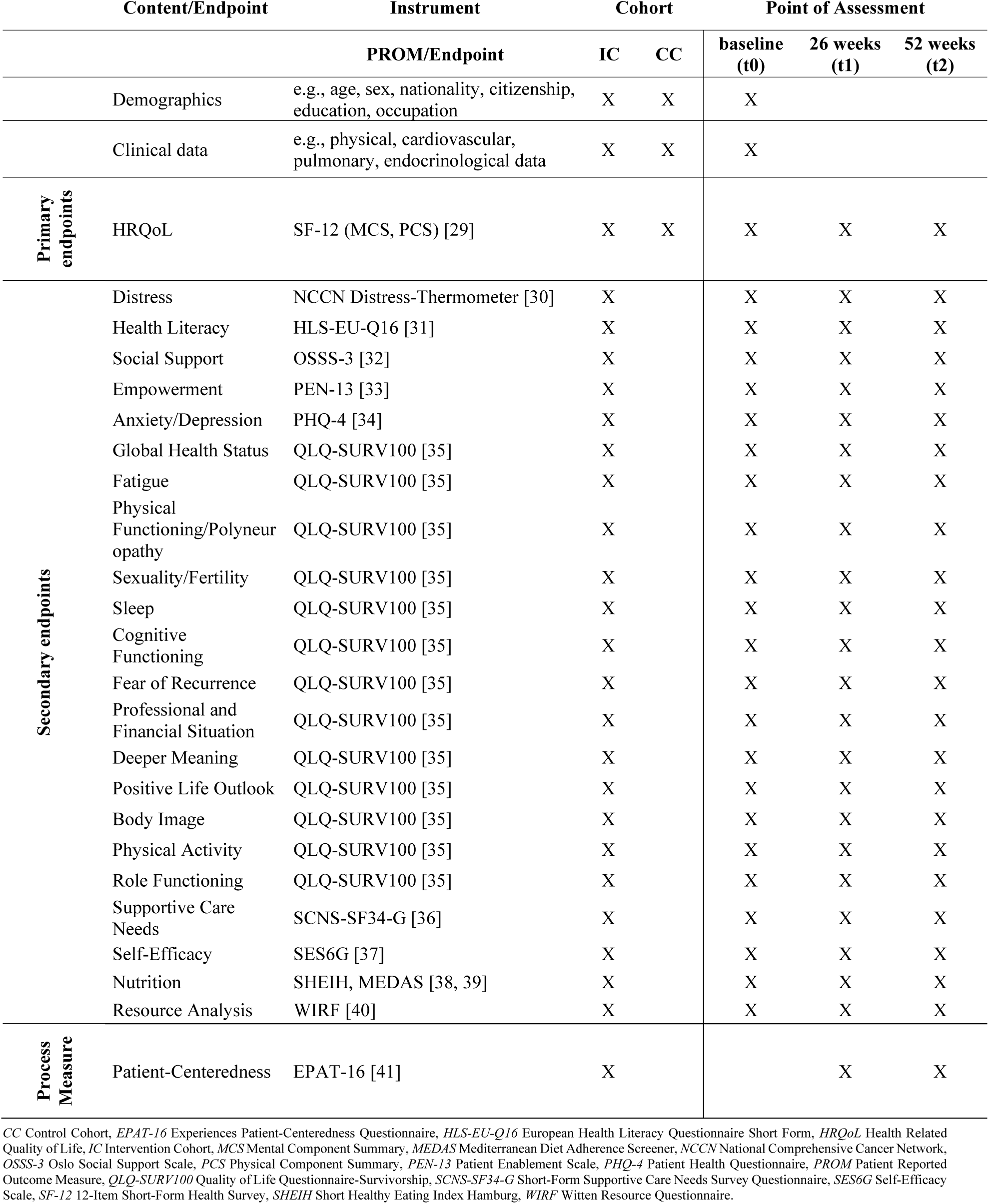
Overview of primary and secondary outcomes, measures and point of assessment in each cohort.

### Recruitment

The program is promoted through several channels, including a project-specific website with a digital flyer available for download and printed flyers distributed to the clinical network, cancer counseling centers, and self-help groups in the metropolitan area of Hamburg. Additionally, the program is presented at clinical meetings across the various departments of the UCC Hamburg network, ensuring awareness and engagement among healthcare professionals. Accordingly, participants will be recruited through multiple pathways: (1)self-referral by cancer survivors; (2) referral via the UCC Hamburg’s clinical network, including private practice oncologists and hospitals in the Hamburg metropolitan region; (3) referral via primary care physicians; (4) referral via self-help groups and patient organizations and (5) referral via cancer counseling centers. After initial registration or referral to the *Hamburg Life After Cancer Program*, participants are contacted for an informed consent telephone call, which is conducted by a team member (HF) using a standardized script. Details on the participant pathway are presented in Figure 2. The control cohort, receiving usual care, will be recruited from routine clinical follow-up care at UKSH.

### Effectiveness Evaluation

For effectiveness evaluation, the selected outcomes presented in Table 1 will be assessed using validated measures at three assessment points: baseline (start of the intervention, t0), 26 weeks (t1), and 52 weeks (t2). The timepoint of assessments for primary and secondary endpoints varies by study group (intervention vs. control) and outcome. We primarily hypothesize, that the *Hamburg Life After Cancer Program* improves HRQoL within the intervention cohort 52 weeks after baseline (t2) compared to baseline (t0). Secondary hypotheses are exploratory and focus on improvement of HRQoL within the intervention cohort 52 weeks after baseline (t2) in comparison to the external control cohort as well as on improvement of various bio-psycho-social outcomes (secondary endpoints, Tab. 1) within the intervention cohort only. For detailed demographic and clinical baseline data assessed in this study see Additional File 1.

### Implementation Evaluation

For the evaluation of the implementation, we apply a mixed-methods approach with qualitative and quantitative data being collected concurrently with program implementation, with a focus on process evaluation. The evaluation framework is guided by the criteria proposed by Proctor et al. [42] and informed by the Consolidated Framework for Implementation Research (CFIR) [43].

Acceptability and feasibility are assessed from both health care professionals’ and cancer survivors’ perspectives through qualitative, semi-structured interviews. The interview guide is structured according to CFIR [43] domains. Interviews are conducted throughout program delivery. Interview-derived feedback will be communicated to the core team as soon as recurring themes emerge or a clear need for action is identified, to inform potential program adaptations. Implementation analysis is further supported by field notes collected continuously throughout the study’s course conducted by the core team to identify implementation barriers and inform potential program adaptations. Feedback derived from field notes is summarized and reported to the core team quarterly.

As further part of the process evaluation, patient-reported experiences of patient-centeredness are assessed using the Experienced Patient-Centeredness Questionnaire (EPAT-16) [41] at t1 and t2. Results from t1 are fed back to the core team in a predefined, stepwise manner to inform potential program adaptations or additional training, complementing the trainings conducted during preparatory study phases (Fig. 1). Initial feedback will be provided after the first 50 completed questionnaires at t1 (corresponding to approximately 10% response rate). Subsequent feedback will be provided at predefined intervals of approximately 20% additional response rate. Furthermore, field notes and feedback derived from the EPAT-16 [41] are systematically reviewed and evaluated during ongoing patient and expert advisory board meetings to facilitate participatory reflection and inform program adaptations throughout ongoing implementation.

Utilization rates of recommendations outlined in the participants’ individual survivorship plan as well as of program components are documented. Additionally, each presentation of the program at clinical conferences, as described in section *Recruitment,* will be documented, including location and presenter. Further, the locations and quantities of distributed flyers will be documented.

### Sample Size

#### Effectiveness Evaluation

The sample size calculation is based on the first hierarchical primary endpoint, the Mental Component Summary (MCS). The second primary endpoint, the Physical Component Summary (PCS), both measured by the 12-Item Short Form Health Survey (SF-12) [29], will be tested hierarchically, conditional on the significance of the first endpoint, without separate sample size calculation.

A total of n=500 participants are planned to be recruited for the intervention cohort over a period of 24 months (Fig. 1), accounting for an anticipated drop-out rate of 20%, resulting in an expected sample of n=400 participants available for analysis at t2. This sample size is primarily determined to ensure adequate precision for estimating the mean change in HRQoL, measured by SF-12 [29], from baseline (t0) to 52 weeks (t2). Specifically, it allows estimation of a two-sided 95% confidence interval with a margin of error of ±1, assuming a standard deviation (*SD*) of 10. The calculations were carried out with PASS 22.0.4 [44] for a confidence interval for paired means.

For the control group, n=250 participants will be recruited, yielding evaluable data from approximately n=200 participants at t2 after accounting for a 20% drop-out rate. For the between-group comparison of change in HRQoL from t0 to t2, these sample sizes allow estimation of a two-sided 95% confidence interval for the mean difference with a margin of error of ±1.7, assuming an *SD* of 10 in both groups. Moreover, with the expected sample size, a standardized mean difference of *d*=0.24 can be detected using an independent-samples t-test with 80% power at a two-sided significance level of α=0.05. The calculations were carried out with PASS 22.0.4 [44] for a confidence interval for the difference between two means and for the two-sample t-test based on an effect size.

#### Implementation Evaluation

For the qualitative interviews evaluating implementation, approximately n=10-15 interviews with health care professionals and n=20-30 interviews with cancer survivors are planned. These numbers represent estimates, since the final sample size for the qualitative analysis will be determined by theoretical saturation.

### Data Analysis

#### Effectiveness Evaluation

A Statistical Analysis Plan will be finalized before data base lock. To evaluate the effectiveness of the intervention within the intervention cohort, changes in the two hierarchical primary endpoints - MCS and PCS scores of the SF-12 [29] - from t0 to t2 will be analyzed using linear mixed-effects models. Adjusted means, 95% confidence intervals and p-values will be reported. Assessment points (t1, t2) and baseline values of the scores will be included as fixed effects, and participants will be treated as random effects. PCS will only be interpreted confirmatory if the MCS change confidence interval does not include zero. Secondary endpoints, including PROMs and standardized scales (Tab. 1), will be analyzed exploratively using analogous modeling strategies appropriate for the scale of each endpoint, with baseline values included as covariates. Pre-specified rules for handling missing items have been defined for each instrument and scale, respectively, as part of the analysis manuals. Missing values will not explicitly be imputed as part of the primary analyses but within a mixed model approach. Multiple imputation will be applied in sensitivity analyses if >5% of primary endpoint data are missing, separately for baseline and follow-up, and only for the intervention group. Comparisons with the external control cohort at UKSH will use the same models with group as a fixed effect, and sensitivity analyses will apply propensity score matching based on age (<40/ ≥40 years), gender (male/female/diverse), tumor entity according to ICD-10 [26] (hematological/oncological), treatment modality (local/systemic/multimodal), and time since diagnosis (<1 year/1-5 years/>5 years). Exploratory mediation analysis is planned, but will be specified in the further course of the study. After approximately half of the participants have been enrolled in either cohort, a planned descriptive analysis will be conducted to assess comparability regarding the following variables: age (<40/≥40 years), gender (male/female/diverse), tumor entity according to ICD-10 [26] (hematological/oncological), treatment modality (local/systemic/multimodal), and time since completion of treatment (<1 year/1-5 years/>5 years). This analysis is intended to ensure cohort comparability and inform further recruitment strategies.

#### Implementation Evaluation

For qualitative data, audio-recorded interviews will be transcribed verbatim. Two study members with experience in qualitative methodology will independently analyze transcripts and field notes imported into MAXQDA (VERBI GmbH, Berlin, Germany) using both inductive and deductive qualitative content analysis [45], where CFIR domains [43] will serve as the a priori coding framework for deductive analysis. Regular meetings with a third researcher will be held to ensure that coding schemes and analyses are comparable. Quantitative, EPAT-16 patient-centeredness data [41] (Tab. 1) will be analyzed descriptively only.

### Data Monitoring

The research team will continuously document data collection and management across all measurement points. Completed questionnaires will be transferred to the clinical documentation system for clinical routine use, which is subject to legal medical documentation requirements. For research purposes, data will be entered into a pseudonymized electronic study database (REDCap) (Vanderbilt University, USA), with only clinical information relevant to the research objectives included in the analysis. Data accuracy will be ensured through double data entry by two independent researchers. Individual patient characteristics can only be identified via a separate linkage list stored securely on a UKE server. Access to both the pseudonymized database and linkage list is restricted to trained intern research personnel, who are responsible for data entry and regular verification of consistency, accuracy, and completeness.

### Patient and Public Involvement

We apply a participatory research approach in collaboration with an *expert advisory panel* and a *patient advisory board,* who accompanies and advises us throughout the entire study - from development and implementation to evaluation of the *Hamburg Life After Cancer Program* (Fig. 1).

The *expert advisory panel*, composed of members with diverse expertise (e.g., complementary medicine, psycho-oncology, self-help, palliative medicine and caregiving research), provides specialized guidance on methodological, clinical, and implementation-related issues and is consulted regularly every six months to support evidence-based decision-making.

The *patient advisory board* consists of a heterogeneous group of cancer survivors and relatives. All members completed a six-month patient ambassador training at the Patient Competence Center North (UCC Hamburg and UKSH), providing foundational knowledge in research, clinical care, and health system. Additional project-specific training (e.g., survivorship, data protection, ethics) is offered as needed.

Principles for effective patient involvement in cancer research [46], consistent with international recommendations [47], guide participation. Core to this approach is the early and active involvement of people affected by cancer in planning and conducting the research, ensuring a comprehensive perspective, alignment with patient needs, and effective collaboration. Trust between stakeholders is essential, and participant heterogeneity is addressed by providing accessible formats and tools. Regular in-person or online meetings occur monthly for the first six months and approximately every six months thereafter to promote dialogue and mutual understanding, supported by clear communication structures and transparent project procedures. Members receive annual compensation for their participation.

During the preparatory study phase (Fig 1.), patient advisory board members actively contribute to finalizing the intervention and study materials, as well as to planning implementation and evaluation (e.g., selection of PROMs). Mid-intervention (t1) EPAT-16 patient-centeredness data [41] as well as feedback derived from fieldnotes and interviews will be jointly reviewed to discuss and identify potential improvements to the program. Following completion of the evaluation, the patient advisory board will be asked to support dissemination efforts, including co-authorship of publications (as in this manuscript, patient advocate OK).

## Discussion

This study addresses a gap in cancer survivorship care and research by developing, implementing, and systematically evaluating a structured, multi-component survivorship program for adult cancer survivors in Germany. Although cancer survivorship has gained international attention, evidence-informed, patient-centered models of survivorship care remain limited in many European health systems [13]. By grounding the *Hamburg Life After Cancer Program* in patient-centered principles and participatory research methods, this study contributes to advancing tailored survivorship care that reflects cancer survivor’s needs and preferences.

We expect improvements in HRQoL along with secondary outcomes such as health literacy, self-management, empowerment, self-efficacy, and further bio-psycho-social outcomes. Strengthening these domains is increasingly recognized as crucial for dealing with late and long-term effects and in supporting cancer survivors in navigating survivorship care structures and daily-life challenges [13, 19, 21]. The generic SF-12 [29] component scores (MCS and PCS) were chosen as primary outcomes as they provide comprehensive assessment of HRQoL across heterogeneous patient groups and care settings. While cancer survivorship-specific instruments such as the Quality of Life Questionnaire Survivorship (QLQ-SURV100) [35] capture detailed survivorship issues, the SF-12 [29] allows broader comparisons, including general population norms, supporting evaluation of the program’s overall impact on HRQoL.

Further, this study applies a pragmatic hybrid type 2 effectiveness-implementation design, as randomization is not considered feasible within the routine care context of the *Hamburg Life After Cancer Program*, which is implemented at the institutional level and integrates multiple components into existing care pathways. While randomized controlled trials are designed to maximize internal validity, their applicability in such settings may be limited, potentially constraining real-world implementation and generalizability [48], particularly for outcomes such as HRQoL, which may be influenced by contextual and care delivery factors [49]. To ensure methodological quality, the study is adequately powered for the primary outcomes and combines a prospective longitudinal design with an external usual-care control cohort. Selection bias cannot be fully excluded, but baseline covariates are examined to ensure comparability and propensity score matching, along with adjusted and sensitivity analyses, is applied to reduce confounding. Residual confounding, particularly from unmeasured factors, may remain. Overall, this approach aims to generate robust evidence on intervention effects on HRQoL under real-world conditions, while transparently acknowledging limitations regarding causal inference.

Beyond evaluating effectiveness, we expect this study to offer insights into key facilitators and barriers to its implementation, using the UCC Hamburg as an exemplary implementation setting, while its design facilitates rapid translation into routine clinical practice. In line with the exploratory nature of this study, broad eligibility criteria are applied and no additional restrictions or regulatory cohort allocation procedures are imposed, allowing for the inclusion of a heterogeneous cancer survivor population and facilitating valid evaluation of feasibility and implementation in routine care.

If demonstrated to be effective and feasible, the *Hamburg Life After Cancer Program* could serve as a blueprint for standardized and structured cancer survivorship care. Ultimately, the findings have the potential to inform national recommendations and contribute to improved long-term HRQoL among cancer survivors.

## Data Availability

Not applicable as no data was generated or analyzed for the production of this study protocol.

## List of abbreviations

HRQoL: Health-Related Quality of Life
PROMs: Patient Reported Outcome Measures
UCC Hamburg: University Cancer Center Hamburg
UKE: University Medical Center Hamburg-Eppendorf
UKSH: University Medical Center Schleswig-Holstein
ICD-10: International Statistical Classification of Diseases and Related Health Problems
CFIR: Consolidated Framework for Implementation Research
EPAT-16: Experienced Patient-Centeredness Questionnaire
MCS: Mental Component Summary
PCS: Physical Component Summary
SF-12: Short Form Health Survey
SD: Standard Deviation
QLQ-SURV-100: Quality of Life Questionnaire-Survivorship.

## Additonal files

File name: Additional file 1.

File format and file extension: Table, Microsoft word .docx.

Title of date: Table A1. Overview of demographic and clinical baseline characteristics.

Description of date: Additional file 1 provides a detailed overview of all demographic data and clinical characteristics, inluding laboratory values, obtained from participants throughout the course of the study.

## Declarations

### Ethics approval and consent to paricipate

The Ethics Committee of the Medical Association Hamburg has approved the study (2025-101485-BO-ff). The study will be carried out in line with the principles of the Declaration of Helsinki. Principles of good scientific practice will be respected. Study participation is voluntary and no foreseeable risks for participants result from the participation in this study. Participants will be fully informed about the aims of the study, data collection, and the use of collected data. Written informed consent will be sought prior to participation. Preserving principles of data sensitivity, data protection, and confidentiality requirements will be met. Written informed consent will be obtained from all individual participants included in the study.

### Consent for publication

Not applicable.

### Availability of data and materials

Not applicable as no data was generated or analyzed for the production of this study protocol.

### Competing interests

HF, JvG, MR, AZ, AKO, AM and HS declare they have no competing interests. FW reports receiving honoraria from Novartis Pharma and having financial relations to Alexion Pharma; no other competing interests are declared. OK reports having received speaker honoraria from AstraZeneca within the past year as part of an employee training program, outside the submitted work; no other competing interests are declared. CB reports receiving speaker honoraria from AOK Germany, Med Update, and Roche Pharma; advisory board fees from AstraZeneca, Bayer Healthcare, BioNTech, Eurobio, Lindis Biotech, Merck Serono, and Oncology Drug Consult CRO; and non-financial interests through advisory, leadership, and board roles in the DGHO (German Society of Hematology and Medical Oncology), the Hamburg Cancer Society, the National Network of German Cancer Centers (DKH), the Northern German Society of Internal Medicine, and the German Cancer Society. AL reports receiving honoraria from AstraZeneca, Bayer, BMS, Böhringer Ingelheim, Grünenthal, Janssen, Lilly, Merck MSD, Novartis, Roche, Servier, Tesaro, Resilience as well as receiving institutional research grant from Amgen, Böhringer Ingelheim and Resilience; no other competing interests are declared. IS reports receiving speaker honoraria from Beiersdorf AG, ClinSol GmbH, Verein für Fort-und Weiterbildung Psychosoziale Onkologie e.V. & Co KG, and Omnicare GmbH within the last 3 years, outside the submitted work; no other competing interests are declared. MS reports receiving honoraria from Astellas, ConPhyMed, Falk, Jazz Pharma and Roche as well as receiving institutional research grant from Abbvie, Amgen, AstraZeneca, BeOne, BMS, Böhringer Ingelheim, GSK and MSD; no other competing interests are declared.

### Funding

This study is funded by the German Cancer Aid (Stiftung Deutsche Krebshilfe), grant number: 70116190.

### Authors’ contributions

MS and IS are the responsible principal investigators of the study. MS, IS, CB, HS and MR contributed to the study conception and design. IS, MS, MR and JvG wrote the grant proposal and obtained funding. HF wrote the first draft of the manuscript. All authors commented on previous versions of the manuscript for important intellectual content. All authors read the final manuscript, gave approval of the version to be published and agreed to be accountable for the work.

## Acknowledgements

We thank the non-author members of the patient advisory board - Ariane Fischer, Fabia von Blücher, Christine Asmussen, Heide Lakemann, Ines Moegling, Silke Vogel, and Souwany Schindler - for their contributions to the development of this study, as well as for their willingness to advise us throughout its entire course. Furthermore, we thank Corinna Bergelt (University Medical Center Greifswald), Inken Hilgendorf (University Medical Center Jena), Christopher Kofahl (University Medical Center Hamburg-Eppendorf), Karin Oechsle (University Medical Center Hamburg-Eppendorf), Oliver Rick (Medical Center Reinhardshöhe), Matthias Rostock (University Medical Center Hamburg-Eppendorf), and Alexander Stein (Hematological and Oncological Outpatient Clinic Eppendorf) for their scientific consultation and continuous guidance as members of the expert advisory panel.

## Notes

### Clinical Trial

Clinical trial registration number: DRKS00035126, German Clinical Trial Register (date of registration: 27 August 2025).

### Author Declarations

The Ethics Committee of the Medical Association Hamburg gave ethical approval for this study (2025-101485-BO-ff).

