## Additional File 1 for "The “Hamburg Life After Cancer Program”: Development, Implementation and Evaluation of a Structured Survivorship Program - Study Protocol of a Hybrid Effectiveness-Implementation Study"

[a] First author

[b] Isabelle Scholl and Marianne Sinn share senior authorship

**Corresponding Author**

Name: Hannah Führes

University Medical Center Hamburg-Eppendorf

Department of Medical Psychology

Martinistr. 52, 20246 Hamburg, Germany

**Additonal File 1**

Table A1. Overview of demographic and clinical baseline characteristics

| | |
|---|---|
| **Demographics** | Age |
| | Sex |
| | Nationality/Citizenship |
| | Migration background |
| | Health Insurance Status |
| | Relationship Status/Marital Status |
| | Number, Age and Sex of Children (*if applicable*) |
| | Education |
| | Occupation/Employment Status |
| | Monthly Income (participant/people involved/living in household) |
| | Degree of disability (*if applicable*) |
| | Care needs/Care service (participant/people living in household) (*if applicable*) |

| | |
|---|---|
| **Clinical Data** | Medical History/Anamnesis (Prior results/findings, comorbidities, tobacco use, cannabis use, alcohol consumption, use of other drugs, current medication) |
| | Oncological Disease (tumor entity ICD-10 diagnosis, time since initial diagnosis, tumor stage) |
| | Physical Examination (height, weight, blood pressure (BP), ECOG performance status) |
| | Laboratory Values (albumin, TSH, free T3 (fT3), free T4 (fT4), triglycerides, HbA1c, LDL cholesterol, HDL cholesterol, creatinine, GFR, GPT (ALT), GOT (AST), gamma-GT (gammaGT), complete blood count with differential (CBC with diff), ferritin, LDH, urine analysis) |
| | Primary Oncological Therapy (start of therapy, end of therapy, therapy within a clinical trial, chemotherapy, high-dose chemotherapy, immunotherapy, radiotherapy, surgery, clinically relevant complications, primary therapy response) |
| | Relapse (yes / no) |
| | Second Primary Tumor (yes / no) |
| | Late Effects / Long-term Effects (fatigue, polyneuropathy, cardiovascular diseases, therapy-associated pain, endocrinopathy, osteopenia / osteoporosis / osteonecrosis, lymphedema, visual impairment, hearing impairment, incontinence, sleep disorders, psychological distress, cognitive impairment, impaired fertility, gastrointestinal complaints) |
| | Current Supportive Care (psycho-oncological support, psychiatric care, self-help group support, social counseling participation, participation / connection to cancer society and its services, participation in rehabilitation) |
| | Additional Information (primary provider for follow-up care/who is currently responsible for follow-up care) |
